# Emergence and fast spread of B.1.1.7 lineage in Lebanon

**DOI:** 10.1101/2021.01.25.21249974

**Authors:** Mahmoud Younes, Kassem Hamze, Hassan Nassar, Mohamad Makki, Maysaa Ghadar, Paul Nguewa, Fadi Abdel Sater

**Affiliations:** Research Department, Beirut Cardiac institute, Old airport road, Beirut, Lebanon; Laboratory of Molecular Biology and Cancer Immunology (Covid 19 Unit),Faculty of Sciences I, Lebanese University, Hadath, Beirut, Lebanon; Research Department, Bahman Hospital, Beirut, Lebanon; University of Navarra, ISTUN Instituto de Salud Tropical, Department of Microbiology and Parasitology. IdiSNA (Navarra Institute for Health Research). c/ Irunlarrea 1, 31008 Pamplona, Navarra. Spain

## Abstract

The severe acute respiratory syndrome coronavirus 2 (SARS-CoV-2) remains a rapid spread emerging disease. Recently, a new variant of this virus called SARS-CoV-2 VOC 202012/01 (or B.1.1.7 lineage), described in the United Kingdom (UK), has become highly prevalent in several countries. Its rate of transmission has been estimated to be greatly higher. B.1.1.7 lineage harbors 23 mutations co-existed for the first time in the same variant. Herein, we are interested only by the deletion mutation ΔH69/ΔV70 in the spike protein.

In the UK they were able to identify the increase of this new variant through the increase in the false negative result for the spike target of a three-target RT-PCR assay from Thermo Fisher Scientific (TaqPath kit). Later, the manufacturer announced that this false negative result is because of the deletion ΔH69/ΔV70 in the area targeted by the TaqPath Kit. Furthermore, The European CDC recommended that the use of this kit help to track the new variant.

Genome sequencing is the gold method to confirm the new variant, but observational studies provide also stronger evidence if similar models are observed in multiple countries, especially when randomized studies are not possible. In Lebanon, the highest number of confirmed cases were reported in first week of 2021. In the present study, we show the emergence and the fast spreading of the new variant in Lebanon and a relationship between SARS-CoV-2 transmission intensity and the frequency of the new variant during the first twelve days of January.

## Introduction

The severe acute respiratory syndrome coronavirus 2 (SARS-CoV-2) was, and remains, the rapid spread emerging disease of the year.^1, 2^ Recently, a new variant of this virus called SARS-CoV-2 VOC 202012/01 (or B.1.1.7) has been described in the United Kingdom (UK). It has become highly prevalent in parts of the UK, and is emerging in other countries. This variant was predicted to potentially be 50 to 70% more rapidly transmissible than other SARS-COV-2 variants circulating in the UK.^3^

The analysis of Sequences showed that the new variant had accumulated a large number of mutations that together caused 17 amino acid changes in the virus’ proteins. This variant was defined by eight mutations in spike protein while the others occurred in other regions of the genome, such as in the *orf1-ab, orf8*, and nucleocapsid (N) genes. One of spike mutations was 69-70del, a deletion in the S1 N-terminal domain NTD, that was found in a patient (Cambridge, U.K.), seemed to allow the virus to evade the immune system.^4^ This mutation increased infectivity of Spike by two fold. While the other mutations in spike protein were deletion 144, N501Y, A570D, D614G, P681H, T716I, S982A and D1118H.

Recently, the European CDC recommended that multi-target RT-PCR assays that included an S gene target affected by the deletions could be used as a signal for the presence of the 69-70del mutation for further investigation and could be helpful to track these mutant variants.^6^

Multi-target RT-PCR tests using S gene regions impacted by the 69-70del mutation are quicker and cheaper than sequencing especially, if sequencing capacity is limited. The Applied Biosystems™ TaqPath™ COVID-19 assay allows the detection of this mutation.^6^ It was shown that in a patient infected with a variant harboring the 69-70del mutation, there will be an S gene “drop out”. The sequencing of diagnostic PCR amplicon products from S gene dropout samples showed that the S gene target contained the 6-nucleotide deletion (21,765–21,770) leading to the Δ69-70 deletion in the middle of the amplicon. Those authors inferred the S gene dropout must be due to a failure of the qPCR probe to bind as a result of the Δ69-70.^7^

Since February, SARS-COV-2 emerged in Lebanon and recently we have observed rapid spread of COVID-19 through the country. On December 21, the first case of the new variant was reported in Lebanon in a traveler on a flight coming from London. The Lebanese government confirmed 226,948 cases of COVID-19 and 1,70512 related deaths in the country in January 12, 2021. The highest amount of confirmed cases was recorded in the first week of 2021. They imposed a countrywide lockdown to combat the spread of COVID-19. In the present study, we show the emergence and the fast spreading of the new variant in Lebanon.

## MATERIALS AND METHODS

### Clinical specimens

Samples were collected in Molecular labs at Bahman Hospital and Beirut Cardiac Institute. All patients or (guardians) provided written and signed informed consents. The study was conducted between December 9, 2020 and January 12, 2021. Nasopharyngeal swab samples were collected from 11657 patients. All PCR tests were performed using Applied Biosystems™ TaqPath™ COVID-19 assay. 2218 of tested patients were reported as positive.

### SARS-CoV-2 rRT-PCR

Viral RNA from Nasopharyngeal swab were extracted, 200 µL of VTM were used for RNA purification. RNA was extracted from the clinical samples on Kingfisher flex purification system Thermo Fisher using MagMAX™ Viral/Pathogen Nucleic Acid Isolation Kit (Thermo fisher). Reactions were performed in a 20-µL final volume reaction containing 5 µL of extracted RNA, rRT-PCR was performed using QuantStudio 5 real-time PCR detection system (Thermo fisher) and TaqPath 2019-nCoV real-time PCR kit (Thermo fisher), which targeted the RdRP, N and Spike genes of SARS-CoV-2. Positive samples were retested by using Kylt^®^ SARS-CoV-2 Complete RTU (AniCon Labor GmbH **Kylt**^®^) which targeted the IP4 and Spike genes.

### RESULTS AND DISCUSSION

In December 9, a local patient was diagnosed as COVID -19 positive patient by PCR using TaqPath 2019-nCoV real-time PCR kit. RDRP and N genes of SARS-COV-2 were detected with a Ct value 18 while S gene was not detected. Two days later, the same profile was also detected in two patients from the same family in our laboratory. The 3 samples were retested using Kylt® SARS-CoV-2 Complete RTU, the results showed a detection of IP4 and Spike target genes. These observations showed that S gene may be affected by the same mutation in these 3 samples. In December 14, the UK health minister first announced that there was a new variant of coronavirus called B.1.1.7 with several mutations in spike protein and other regions of the genome. The major mutation in spike gene was the deletion of 6-nucleotide (21,765–21,770) leading to the Δ69-70 deletion. However, between 12 and 24 December we detected the S gene dropout in 10 other patients. In December 24, Thermofisher announced that the Applied Biosystems™ TaqPath™ COVID-19 assay, using S gene regions impacted by the 69-70del mutation (found in the middle of the target amplicon), could be used as a signal for the presence of this mutation. These findings led us to suggest that our samples with S gene dropout could be the B.1.1.7 variant that emerged in Lebanon at least from December 9. Furthermore, it was reported an increase in the number of patients infected with this new variant from December 23. While the first confirmed case infected by the new variant was announced by the Lebanese government, the patient was a passenger arriving from London on December 22. Furthermore, we recorded 498 confirmed cases during the last week of December, among them 52 cases were infected by the new variant. However, in the first week of 2021, the number of cases infected with the new variant increased dramatically. In January 1^st^, 16% of positive cases tested in our laboratory was infected by the new variant. Over a period of 12 days, the number of patients infected with this new variant has dramatically increased reaching approximately 60% of positive cases (figure 1). The new variant is now the focus of intense debate and analysis, and countries worldwide are scouring their own databases to see if it is circulating within their borders. So far, only a few cases of the UK’s new variant have been reported globally. Cases have been found in Iceland, Denmark, the Netherlands, Italy and Belgium and recently in France. In the present study, and during this ongoing COVID-19 pandemic and the emergence of multiple viral variants, we illustrate the emergence and the fast spreading of the new variant in Lebanon from December 9. Such an analysis will be carried to measure the frequency of mutations in our community; then, to determine its local transmission in Lebanon. Based on its spreading, we believe that the new variant will be the most common variant around the world.

**Figure 1.**
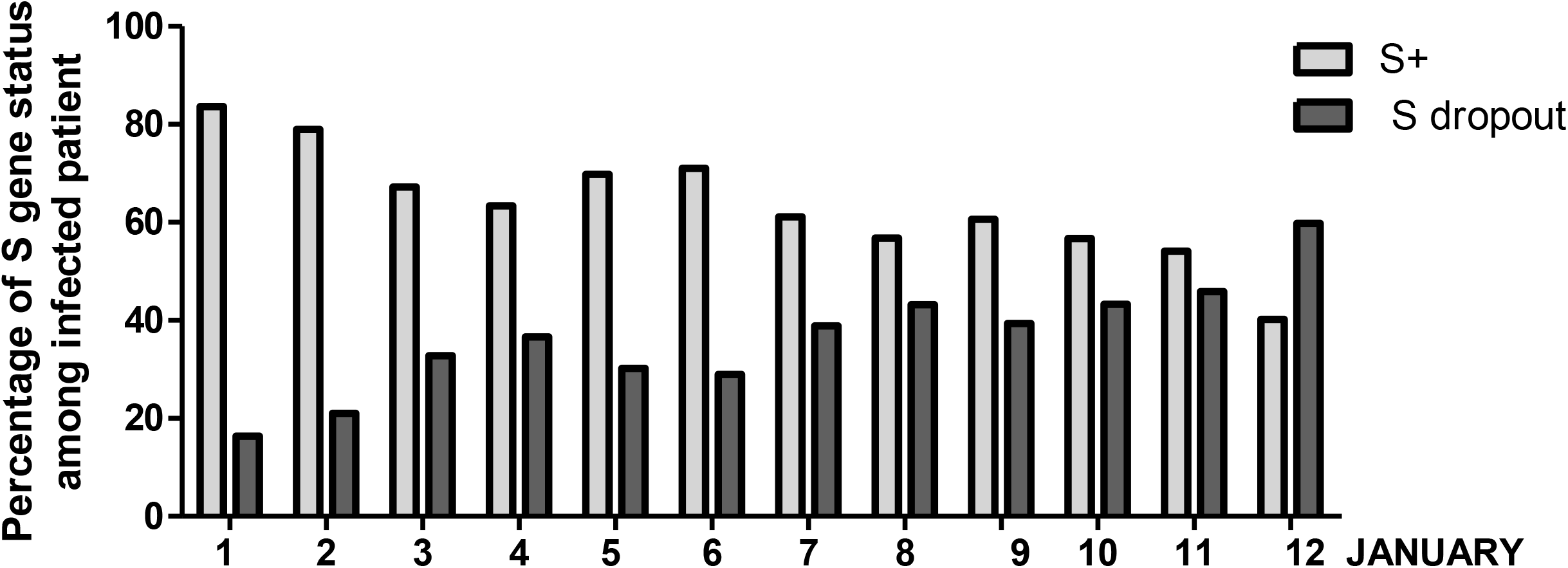
Distribution of infected patients according to the Δ69-70 deletion over a period of 9 days. Of the 5,247 patients tested in January, 1,211 were positive, of which 475 were infected with the new variant. RT-PCR was performed using TaqPath 2019-nCoV real-time PCR kit (Thermo fisher), which targeted the RdRP, N and Spike genes of SARS-CoV-2. All PCR tests were performed using Applied Biosystems™ TaqPath™ COVID-19 assay. S+ : spike detection, S dropout: spike not detected

## Data Availability

available upon request

## ACKNOWLEDGEMENTS

PN thanks Fundación La Caixa (LCF/PR/PR13/11080005), Fundación Caja Navarra, Fundación Roviralta, Ubesol, Inversiones Garcilaso de la Vega, COST Actions CA18217 and CA18218, and EU Project uncover (Grant/Award Number: 101016216) for their support. FAS and KZ thank the Lebanese University its support.

